# Molecular detection of SARS-CoV-2 using a reagent-free approach

**DOI:** 10.1101/2020.04.28.20083626

**Authors:** Ronan Calvez, Andrew Taylor, Leo Calvo-Bado, Colin Fink

## Abstract

Shortage of reagents and consumables required for the extraction and molecular detection of SARS-CoV-2 RNA in respiratory samples has led many laboratories to investigate alternative approaches for sample preparation. Fomsgaard *et al* 2020^5^ recently presented results using heat-processing of respiratory samples prior to RT-qPCR as an economical method enabling an extremely fast streamlining of the processes at virtually no cost.

Here, we present our results using this method and highlight some major pitfalls that diagnostics laboratories should be aware of before proceeding with this technique. We first investigated various treatments using different temperatures, incubation times and sample volumes based on the above study to optimise the heat-treatment conditions. Although the initial data confirmed the published results, further investigations revealed unexpected inhibitory properties of some commonly used virus transport media (VTMs) on some commercially available RT-qPCR mixes, emphasising the critical importance of a thorough validation process to determine the most adapted reagents to be used depending on the sample types to be tested.

In conclusion, although the method works, with very consistent Ct values and an excellent sensitivity when compared to a conventional RNA extraction method, it is critical to include an internal control to check each sample for potential inhibition.

## Introduction

A novel severe acute respiratory syndrome coronavirus-2 (SARS-CoV-2) recently emerged in the Wuhan province of China in December 2019 and is now recognised as the cause of the present human coronavirus pandemic (Covid-19)^1,2,3^. Due to the unprecedented high demand in SARS-CoV-2 testing required to identify and isolate infected individuals, many laboratories are now facing shortages in reagents and consumables required for nucleic acid (NA) extraction. This has led us to investigate alternative sample processing methods that would remove the requirements for NA extraction^4,5,6,7^ and allow rapid diagnosis.

Examining RNA in respiratory samples directly would aid high-throughput screening and sample preparation at low cost, Fomsgaard *et al* 2020 recently presented results using heat-processing of respiratory samples prior to RT-qPCR as a very attractive method enabling an extremely fast streamlining of the processes.

Here, we provide some recommendations based on our results and highlight some major pitfalls that diagnostics laboratories should be aware for processing patient samples for the diagnostics of SARS-CoV-2.

## Methods

We investigated various treatments using different temperatures, incubation times and sample volumes based on Fomsgaard *et al.2020* methodology using heat-processing of respiratory samples prior to RT-qPCR for the detection of SARS-CoV-2.

To test this methodology, 25μL or 40 μL volumes of samples were added to a 96 well PCR plate, sealed and heat-treated at either 95°C or 98°C for 5 minutes on a UKAS-calibrated PCR block. At the end of the incubation, the samples were transferred to another thermoblock set at +4°C for at least 5min. Twenty μl of each lysate was then transferred to a 96-well PCR plate containing various commercially available 1-step RT-qPCR mix for SARS-CoV-2 detection. The assay was then performed and the Ct values were recorded for each sample.

SARS-Cov-2-positive (including samples with a very low viral load) and SARS-Cov-2-negative respiratory samples previously tested in our laboratory using our reference N gene assay (see below) were reassessed. Samples were chosen to encompass some of the common swab sample types received at Micropathology Ltd for SARS-Cov-2 testing (swabs re-suspended in their original transport medium and dry swabs resuspended upon reception at Micropathology Ltd in 500 μL 0.1% Igepal CA-630 (Sigma-Aldrich)). The samples used ranged from less than 1,000 (Ct<37) to greater than 100,000 (Ct<30) SARS-Cov-2 RNA copies/mL as determined by our conventional extraction and PCR protocol.

Our conventional SARS-Cov-2 assay included NA extraction using the Maxwell® HT 96 NA extraction kit (Promega Corp) with automated extraction on the KingFisher FLEX platform (Thermo Fisher Scientific Inc). SARS-CoV-2 detection (2019-nCoV_N1 assay derived from the CDC.gov website^8^) was performed using the ABI TaqMan® Fast Virus 1-Step RT-qPCR kit (#4444436, Thermo Fisher Scientific Inc). The lower limit of detection was established at 364 CARS-CoV-2 RNA copies/mL by ddPCR (QX200 from Bio-Rad).

The baculovirus *Adoxophyes orana* granulovirus (AoGV) is an insect DNA virus routinely used at Micropathology Ltd as an extraction and PCR internal control. In routine diagnostics, the whole virus is spiked at a fixed concentration in the lysis buffer prior to sample extraction and detected using an in-house TaqMan hydrolysis probe-based assay targeting the granulin gene of AoGV. In the present study, extracted AoGV DNA was mixed with the sample prior to heat-treatment and was used as a nucleic acid degradation and PCR inhibition control by comparing its level following heat-treatment to its level in non-heat-treated samples.

## Results

### Optimisation of the heat-treatment method and inhibitory properties of UTM

In a preliminary study to evaluate the best assay conditions, a range of SARS-CoV-2-positive and negative samples was selected and heat-treated at different temperatures and for different times. As shown in Table 1, all the dry swab samples (resuspended in 0.1% Igepal at Micropathology Ltd) and the green-cap swab samples (MWE, resuspended in ∑-Virocult® transport medium at source) tested were all correctly detected using a 5min incubation at 95°C followed by a 4°C-incubation for at least 5min. These conditions yielded a sensitivity and specificity similar to the ones obtained using our conventional assay (with consistent Ct values across both methods), supporting the previously published data using a similar method^5^. Shorter (2min) and longer (10min and 20min) incubation times at these temperatures, or at 90°C, did not improve sensitivity (not shown).

**Table 1:**
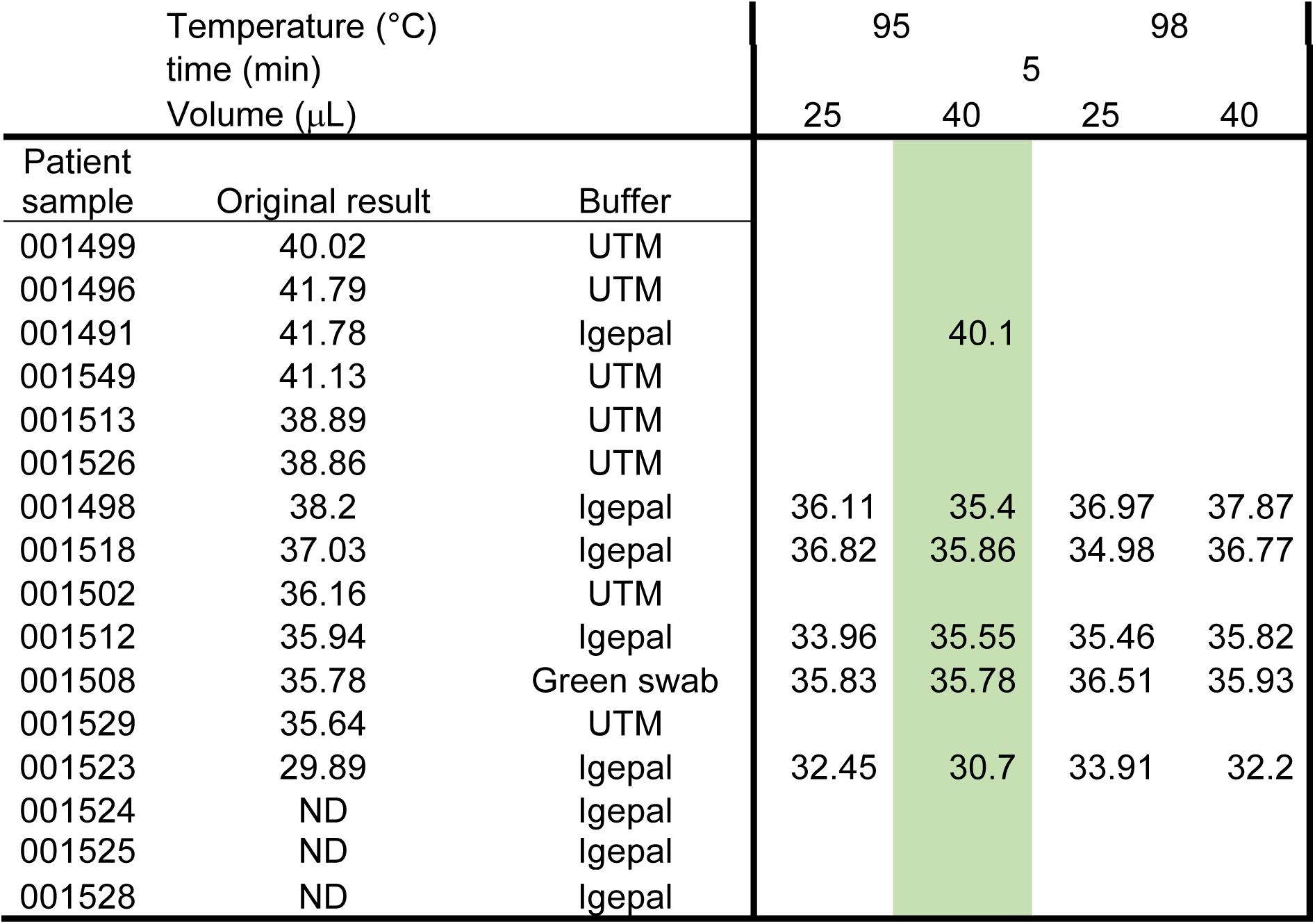
Optimisation of heat-induced sample preparation for SARS-CoV-2 testing. A range of SARS-CoV-2-positive and -negative samples, originally tested using our in-house assay, were subject to heat-treatment using different temperatures and sample volumes. Each heat-treated sample was then subject to SARS-CoV-2 testing using our in-house assay derived from the CDC resource website (targeting the N-gene and using the TaqMan® Fast Virus 1-Step RT-qPCR kit from ABI). The original results were obtained following nucleic acid extraction (Custom Promega Maxwell® HT DNA kit) and purification on a KingFisher platform (ThermoFisher) and are expressed as a Ct value. The buffers correspond to the buffers used in various swab tubes. Igepal (0.1%) is the buffer used at Micropathology Ltd to resuspend dry swabs. Green swabs: ∑-Virocult® transport medium. UTM: universal transport medium used in red-capped COPAN swabs. ND: Not Detected. Highlighted in green are the conditions that were chosen for the rest of the study.

| Temperature (°C)<br>time (min)<br>Volume (μL) |  |  | 95 |  | 98 |  |
| --- | --- | --- | --- | --- | --- | --- |
|  |  |  | 5 |  |  |  |
|  |  |  | 25 | 40 | 25 | 40 |
| Patient sample | Original result | Buffer |  |  |  |  |
| 001499 | 40.02 | UTM |  |  |  |  |
| 001496 | 41.79 | UTM |  |  |  |  |
| 001491 | 41.78 | Igepal |  | 40.1 |  |  |
| 001549 | 41.13 | UTM |  |  |  |  |
| 001513 | 38.89 | UTM |  |  |  |  |
| 001526 | 38.86 | UTM |  |  |  |  |
| 001498 | 38.2 | Igepal | 36.11 | 35.4 | 36.97 | 37.87 |
| 001518 | 37.03 | Igepal | 36.82 | 35.86 | 34.98 | 36.77 |
| 001502 | 36.16 | UTM |  |  |  |  |
| 001512 | 35.94 | Igepal | 33.96 | 35.55 | 35.46 | 35.82 |
| 001508 | 35.78 | Green swab | 35.83 | 35.78 | 36.51 | 35.93 |
| 001529 | 35.64 | UTM |  |  |  |  |
| 001523 | 29.89 | Igepal | 32.45 | 30.7 | 33.91 | 32.2 |
| 001524 | ND | Igepal |  |  |  |  |
| 001525 | ND | Igepal |  |  |  |  |
| 001528 | ND | Igepal |  |  |  |  |

However, all the red-cap tubes for swab samples tested (Copan swabs resuspended in UTM-RT medium, Copan Diagnostics) failed to produce any detectable signal despite a significant viral load (Ct>37). Original results reported in our laboratory were obtained using our conventional protocol (described in the methods section) using the ABI TaqMan® Fast Virus 1-Step RT-qPCR kit.

### Performance of two Meridian Bioscience RT-qPCR mixes

In order to understand the failure of the heat-treatment prior RT-qPCR detection of SARS-CoV-2 in the samples resuspended in UTM, we investigated two additional RT-qPCR kits from another manufacturer using another range of SARS-CoV-2-positive patient samples previously detected using our in-house assay. The three commercially available one-step RT-qPCR kits were tested following a 5min heat-induced treatment at 95°C in 40μL volume: the ABI TaqMan® Fast Virus 1-Step RT-qPCR, the Low LOD One-Step RT-qPCR (Meridian Bioscience #MDX025) and the Fast One-Step RT-qPCR (Meridian Bioscience #MDX032) kits. All kits were used according to the manufacturer’s instructions.

As shown in table 2 below, the ABI TaqMan® One-Step RT-qPCR mix systematically failed to detect SARS-CoV-2 in swab samples resuspended in UTM (COPAN swabs) even in the presence of viral loads greater than 1×10^6^ copies/mL (003850 and 003862). In contrast, the other two RT-PCR mixes tested (Meridian Bioscience) allowed the reliable detection of SARS-Cov-2 even at very low viral loads (Ct<37) in all swab samples tested. This strongly suggested two possibilities: the presence of RT and/or PCR inhibitors, or the degradation of the RNA during the heating process in samples resuspended in UTM. Interestingly, a range of non-swab samples (NPA) received from a different NHS trust failed to allow the successful amplification of SARS-Cov-2 in any of the three positive samples tested positive using our reference protocol (006223–006225) even in the presence of viral load >100,000 copies/mL (Ct 30).

**Table 2:**
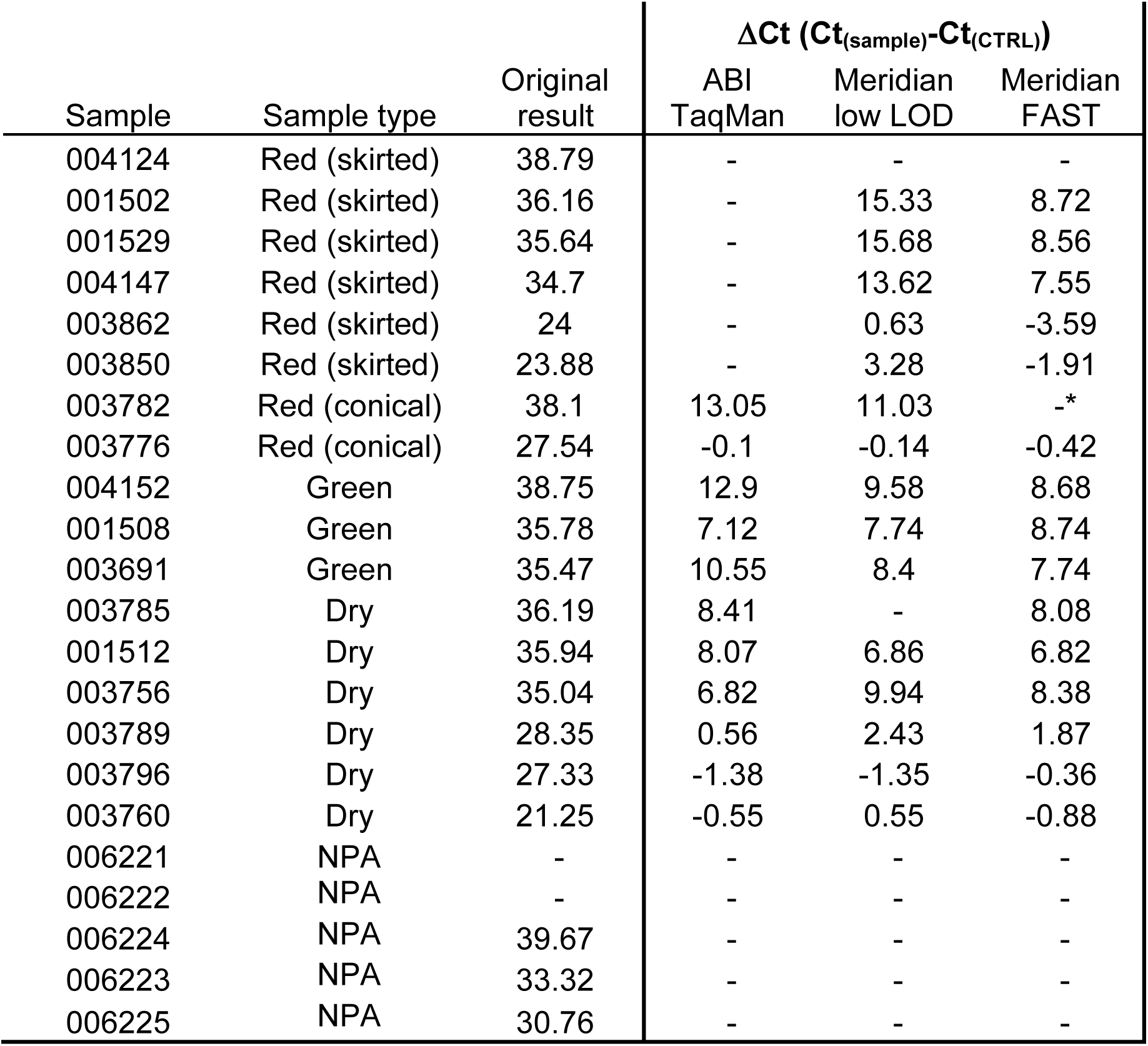
Assessment of transport medium on SARS-CoV-2 detection. Samples that tested positive for SARS-CoV-2 on our in-house assay following nucleic acid extraction (as described in Table 1), were retested using various RT-qPCR mixes following a 5min heat-treatment at 95°C and 5min at +4°C. Ct values from the original results (KF/Promega extraction and ABI TaqMan® Fast Virus 1-Step RT-qPCR) are indicated in the “original results” column. Since the noise band could not be placed at the same level for all samples, results are expressed as the difference between the measured Ct value and the positive control Ct value. Red skirted swab: COPAN swab with UTM-RT for viruses, Chlamydia, Mycoplasma and Ureaplasma. Red conical swab: REMEL M4RT swabs with VTM for transport of viruses and Chlamydia. Green swab: ∑-Virocult swab for transport of viruses. Dry: dry swabs resuspended in 0.1% Igepal. NPA: nasopharyngeal aspirate. “-”: Not detected. *: SARS-CoV-2-positive (Ct 39.63) upon re-test.

### Evidence for PCR inhibitors in respiratory samples

To determine whether the failure of all PCR kits to detect SARS-CoV-2 in the NPA samples was caused by PCR inhibitors, an aliquot of each heat-treated sample was mixed with extracted DNA from a baculovirus (AoGV) suspension. AoGV was then amplified using our in-house assay (see Methods). As shown in figure 1, amplification was successful in two out of 5 of the NPA tested (006222 and 006223), whereas there was a very significant decrease in one of them (006221) and no amplification at all in the two samples where we also failed to detect high concentrations of SARS-CoV-2 (006224 and 006225). These results strongly suggest the presence of PCR inhibitors in some heat-treated samples leading to the generation of false negative results. Our AoGV assay did not reveal the presence of PCR inhibitors in any of the UTM samples tested, suggesting that the ABI RT-qPCR mix is more sensitive to compounds present in the UTM.

**Figure 1:**
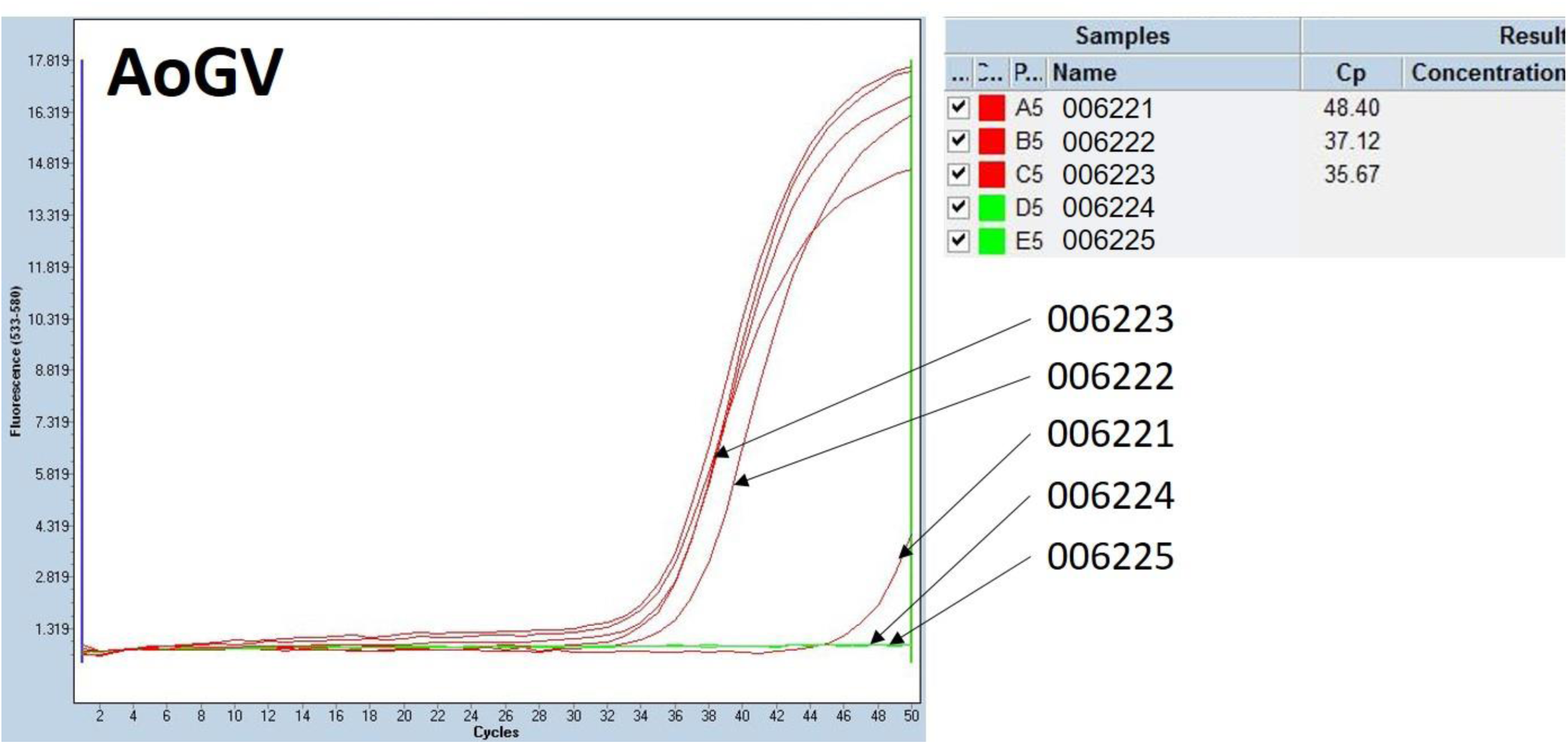
Evidence for PCR inhibition in NPA samples. The 5 NPA samples tested in table 2 were spiked with extracted baculovirus (AoGV) and tested for the presence of AoGV in a TaqMan assay. Samples 006223, 006224 and 006225 were originally tested positive for SARS-CoV-2 using our conventional protocol with Ct of 33.32, 39.67 and 30.76, respectively.

### Sensitivity of the heat-treatment method

To assess the sensitivity of the procedure, 68 patient samples containing weak to intermediate viral loads of SARS-CoV-2 (mean Ct 34.87 ± 4.80 cycles), as assessed by our conventional assay (see Methods), were selected. These samples included all the swab samples types routinely received at Micropathology Ltd for SARS-CoV-2 testing. These samples were recovered from storage (−20°C), thawed, and tested using the methodology described above: 5min at 95°C followed by >5min at 4°C with detection using the Meridian Bioscience Fast 1-Step RT-qPCR kit (N gene). The same samples were also re-tested in parallel using our conventional assay with NA extraction to assess for potential RNA degradation during storage. A proportion (9/68) of the samples that initially tested positive for SARS-CoV-2 on the fresh samples were not detected using both methods, suggesting a detrimental effect of the freeze-thawing cycle on RNA recovery. All of these nine samples contained a sub limit of detection viral load (Ct=38.91 ± 0.93 cycles)

As summarised in the table below SARS-CoV-2 detection performed directly from heat-treated patient samples allowed the successful detection of 73% of the samples that tested positive following re-extraction. All the sixteen heat-treated samples where SARS-CoV-2 amplification was unsuccessful contained a very low viral load of SARS-CoV-2 (mean Ct 40.16 ± 0.98 cycles), whereas all the intermediate/strong positive samples (Ct<37) were correctly detected using both methods. Please note, the sensitivity of the heat-treatment process calculated (73%) does not reflect the true sensitivity of the method since the majority of the strongly positive SARS-CoV-2-samples (Ct<26) were excluded from this cohort of patient samples used in table 3. Multiple parallel diagnostics runs using non-screened samples (n>350) revealed an actual sensitivity of 95.2% after removal of the samples showing signs of PCR inhibition using our AoGV internal control (data not shown).

**Table 3:** Evaluation of the sensitivity of the heat-treatment method. A total of 68 samples that were initially tested positive (mean Ct 34.87) on the conventional assay were thawed and retested for SARS-CoV-2 following re-extraction using the conventional assay (left column) or following heat-treatment (right column). All the non-detected samples were weak positive with an average Ct value of 38.91 ± 0.93.

|  | Reextraction +<br>RT-PCR | Heat-Treatment +<br>RT-PCR |
| --- | --- | --- |
| Total positive | 59 | 43 |
| Average Ct | 37.03 | 36.25 |
| SD Ct | 4.22 | 4.74 |
| % positive | 100.0% | 72.9% |

### Parallel assessment of other commercially available RT-qPCR kits

Using the cohort of patient samples from Table 3, 4 additional commercially available RT-PCR mixes were evaluated. As summarised in Table 4, apart from the Meridian Bioscience kits, all the RT-PCR kits tested failed to detect SARS-CoV-2 in samples re-suspended in the UTM-RT medium used in the COPAN swabs, suggesting the presence of inhibitory factors in this transport medium. In addition, the ABI TaqMan® Fast Virus kit also failed to allow the detection of SARS-CoV-2 in samples re-suspended in the VTM medium used in the Cepheid Xpert® swabs. These results indicate that various transport media may contain specific RT and/or PCR inhibitors that may specifically interfere with the performance of some particular RT-PCR kits, emphasising the essential importance of initial assay condition assessment by each laboratory before proceeding with this technique in order to determine potentially inhibitory sample types.

**Table 4:** Evaluation of the sensitivity of different commercially available RT-PCR kits in different swab sample types. A range of different swab samples previously shown to contain low to intermediate levels of SARS-CoV-2 were retested following heat-treatment (5min at 95°C). Meridian Bioscience FAST 1-Step (#MDX032), Meridian Bioscience Low LOD 1-Step (#MDX025), ABI TaqMan® Fast Virus 1-Step ((#4444436), Quantabio qScript XLT ToughMix® (#95132–100) and Promega GoTaq® 1 -Step (#A6020) RT-qPCR mixes.

|  | Meridian<br>FAST | Meridian<br>LowLOD | ABI TaqMan<br>Fast Virus | Quantabio<br>ToughMix | Promega<br>GoTaq |
| --- | --- | --- | --- | --- | --- |
| Remel swab (M4-RT VTM) | + | + | + | + | + |
| MWE swab<br>(S-Virocult) | + | + | + | + | + |
| Dry swab<br>(Igepal) | + | + | + | + | + |
| Cepheid Xpert®<br>swab (VTM) | + | + | - | + | + |
| Copan swab<br>(UTM-RT) | + | + | - | - | - |
| **Overall detection**<br>**rate** | 72.9% | 69.5% | 33.9% | 33.9% | 55.9% |

### Correlation between heat-treatment and NA extraction protocols

Recent reports using a similar approach (70°C for 10min) suggested a very significant increase in the Ct values (6.1 ± 1.6 cycles) following heat-treatment. To determine the effect of our heat-treatment protocol on patient results, the Ct values of all the positive samples shown in Table 3 were compared. As shown, in figure 2, the correlation between the two assays was excellent (correlation coefficient:1.017) and the Ct values measured using both methods were very consistent, suggesting that, although the heat-treatment slightly affects sensitivity (Table 3), the quantitative results obtained using both methods are highly comparable.

**Figure 2:**
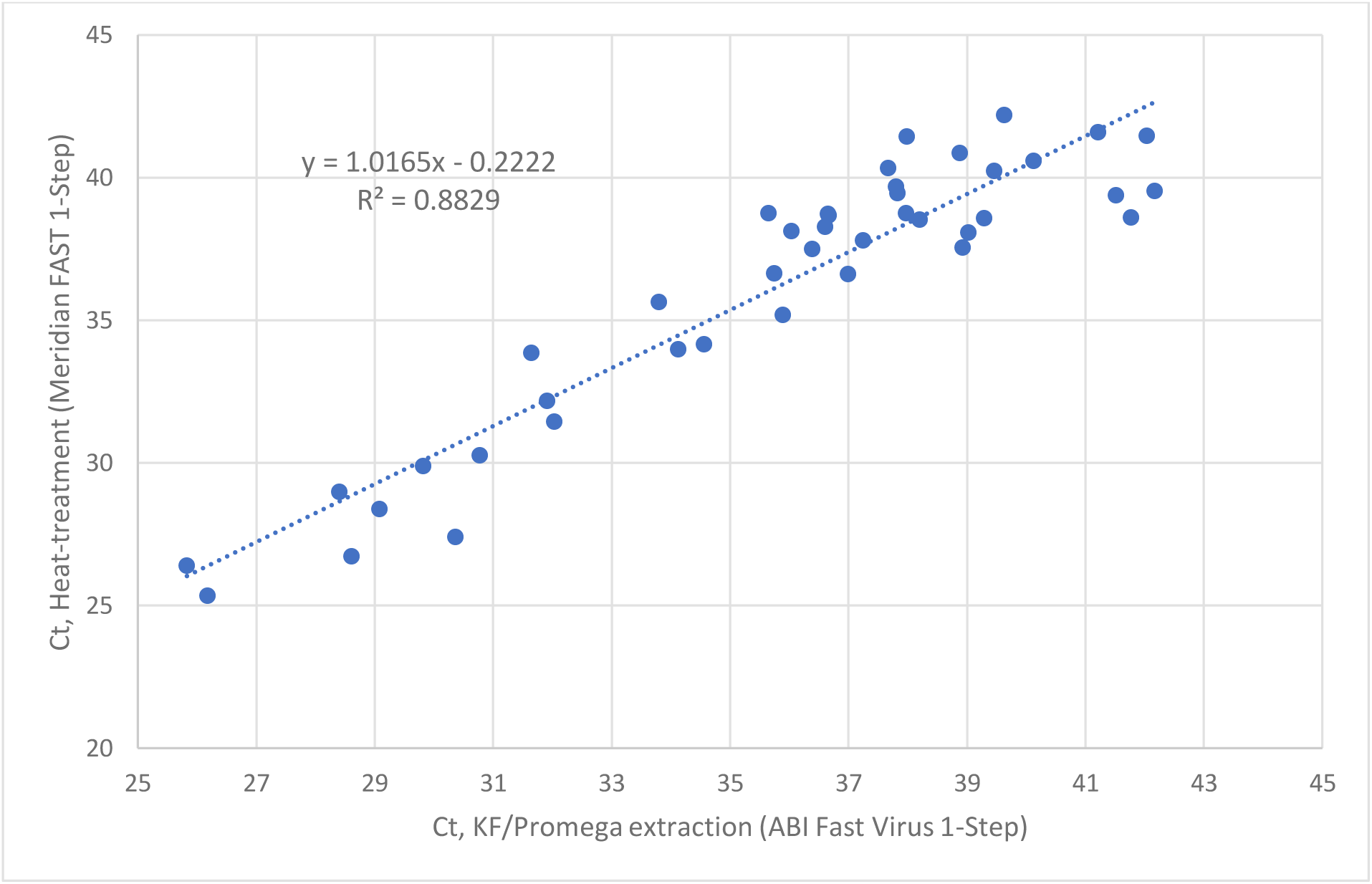
Correlation between the conventional extraction/detection method and the direct detection from heat-treated samples. The Ct values obtained for the samples positive for SARS-CoV-2 using both methods (see Table 3) were plotted against each other.

## Concluding remarks

Although heat-treatment of respiratory samples prior to RT-qPCR showed an attractive methodology compared to a conventional nucleic acid extraction method, the addition of an internal control is critical for quality control and successful detection of this virus if present in the patient samples.

Our results demonstrate and verify the reproducibility and the technical feasibility of a “reagent-free” extraction process which resulted in a detection sensitivity (95.2%) in the same range as others^5^ for most of the samples tested. In addition, we also found a very high correlation between the two methods in terms of Ct values. The success of SARS-CoV-2 diagnostic using this methodology, however, appeared strongly dependent on the properties and robustness of the RT-PCR commercial mix used since some virus transport media (eg UTM from COPAN swabs) appeared incompatible with subsequent SARS-CoV-2 detection in some mixes. In addition, some non-swab samples (eg NPA), showed PCR inhibitory properties rendering them incompatible with all RT-PCR mixes evaluated in this study. Although we did not use the same RT-PCR reagents used by AS Fomsgaard et al, we also recommend a thorough study of all sample types tested should be performed prior to performing direct RT-PCR detection from heat-treated samples.

Although the method presented here offers several undeniable advantages in terms of time and cost savings, our results advise great caution when using such a procedure. The use of internal RT and PCR controls is critical, together with an alternative conventional nucleic acid procedure to process the samples that would fail such internal controls, in order to avoid false-negative results.

## Data Availability

All the experiments and results presented here were performed at Micropathology Ltd.

## Acknowledgement

The authors gratefully acknowledge Dr Mark Atkins for critical review of the manuscript, the staff at Micropathology Ltd especially Dr Oliver Smith, Dr Edward Sumner and Dr Paul Scott for their scientific inputs, as well as collaborators at the University of Warwick, especially Dr David Roper and Dr John Walsh for their support and scientific contributions.

## Ethical statement

This study was performed at Micropathology Ltd (University of Warwick Science Park, Coventry, UK) in April 2020. Patient samples were de-identified and were not considered Human Subjects Research due to the quality improvement and public health intent of the work.

